# Estimation of the true infection rate and infection fatality rate of coronavirus disease 2019 in each country

**DOI:** 10.1101/2020.05.13.20101071

**Authors:** Masahiro Sonoo, Takamichi Kanbayashi, Takayoshi Shimohata, Masahito Kobayashi, Masashi Idogawa, Hideyuki Hayashi

## Abstract

The True Infection Rate (TIR) in the whole population of each country and the Infection Fatality Rate (IFR) for coronavirus disease 2019 (COVID-19) are unknown. We devised a simple method to infer TIR and IFR based on the open data. The estimated TIR was compared with local antibody surveys. Estimated IFR took on a wide range of values up to 10%. The importance of the attenuation of the viral virulence is emphasized.

Coronavirus disease 2019 (COVID-19) pandemic is a worldwide peril. The True Infection Rate (TIR) of COVID-19 in the whole population of each country and the Infection Fatality Rate (IFR) would be important parameters for guiding the strategy of public health, although they are unknown yet. We devised a simple method to infer TIR and IFR based on the open data.

## Data acquisition and analysis methods

We picked up the data at a website 5 times from April 10th to June 13th, 2020, approximately every 2 weeks [1]. Countries or regions (hereafter called just countries) having more than 1000 cases on April 10th were included. The prevalence rate of polymerase chain reaction (PCR) tests among the population (Examination Rate; ER) and the positive rate of PCR tests (Infection Rate; IR) were calculated. The trajectory of each country was drawn over the IR vs. ER plot (logarithmic scales for both). We hypothesized that IR and ER are negatively correlated for a specific country because PCR examinations will be restricted to cases with strong suspicion while ER is low, whereas it will be expanded to general population as ER increases. Then, the TIR of a country can be estimated as the IR value at 100% ER by extrapolating the regression line. The ratio of the infected persons in the population to the already identified cases was calculated and named True/Identified Case Ratio (TICR). IFR was estimated using the number of total deaths on June 13th at the same website [1]. All statistical calculation was done using Microsoft Excel for Macintosh. For each parameter, 95% confidence interval (CI) was calculated using the 95% CI of the slope value of the linear regression line.

## Trajectories of individual countries over the IR vs. ER plot

Included were 64 countries. Trajectories of representative countries are shown in Figure 1. Many countries including most European, Oceanian, and East Asian countries presented with downward-sloping straight lines, which were parallel to each other. Other countries showed upward-sloping or fluctuating lines. An upward line indicates that IR increases despite expansion of the PCR tests, i.e. the infection has been still spreading during the investigation period. Our estimate would not be applicable to these countries since TIR is thought to have also been greatly increasing during the period. In contrast, a downward straight line indicates that our hypothesis on the IR-ER relationship is valid for such a country. Close inspection of individual countries revealed that the first point (April 10th) was shifted downward from the line extended from the latter 4 points in several countries (Belgium, UK, USA, Denmark, and Japan). This is probably due to insufficient containment in these countries until middle of April. Because of this, we decided to calculate the regression line from the latter 4 points.

**Figure 1.**
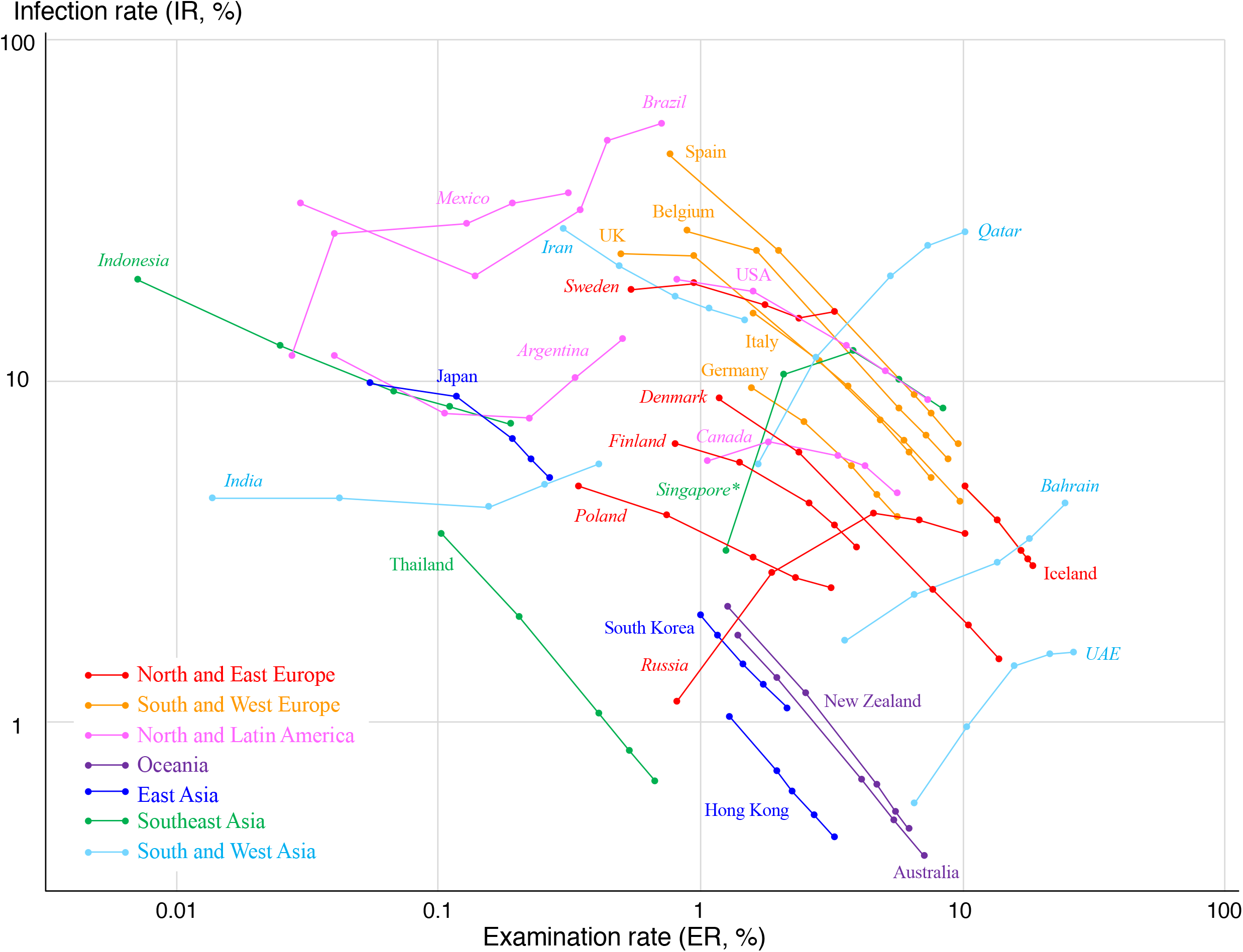
Trajectory of representative countries on the IR vs. ER plot. Countries are color-coded by the region in the world. The names of the countries are also indicated. Italic names mean that the TIR was not estimated because of low correlation. *Singapore*\*: Latter 3 time points form a straight line and therefore TIR was calculated from these 3 points. ER, examination ratio; IR, infection ratio; TIR, true infection ratio.

## Estimates of TIR and IFR

The correlation coefficients and the slope values of the regression line for all investigated countries are listed in Table 1, together with estimated TIR, TICR, and IFR values and their 95% confidence interval (CI). The correlations were often very high, and we confined the estimation to countries having a correlation coefficient between -0.99 and -1. The countries are sorted by the ascending order of the slope value in Table 1. Countries having a slope value close to -1, such as New Zealand, Iceland, or Thailand, are especially noticeable. The slope value of -1 indicates that the number of infected persons remained the same despite expansion of the PCR tests and therefore no patients probably exist in the unexamined population, i.e. the containment has been completely successful. In our preceding study, we defined the containment ratio (CR) as the latest value of the new cases during 7 days divided by its highest value, representing the success of containment [2,3]. CR values are also shown in Table 1. The slope value showed a good correlation (r = 0.784) with CR. Many parallel downward-sloping lines in Figure 1 reflects the wide variety of TIR among countries that has achieved relatively good containment: TIR is generally higher in European countries than in East/Southeast Asian and Oceanian countries. TICR values were usually between 1 and 2 for these countries, especially for European countries, which means that there are not so many infected persons in the unexamined general population.

**Table 1.**
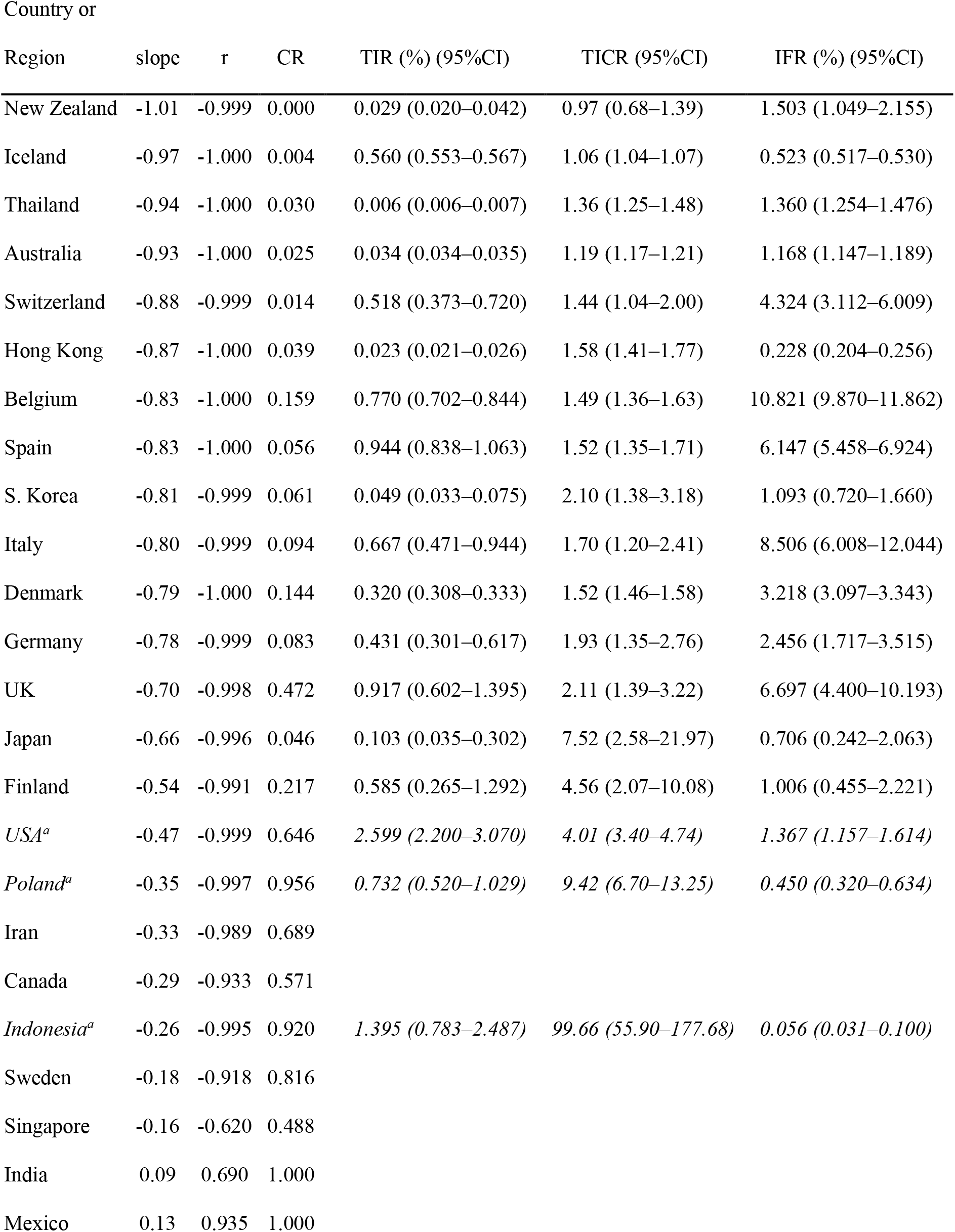
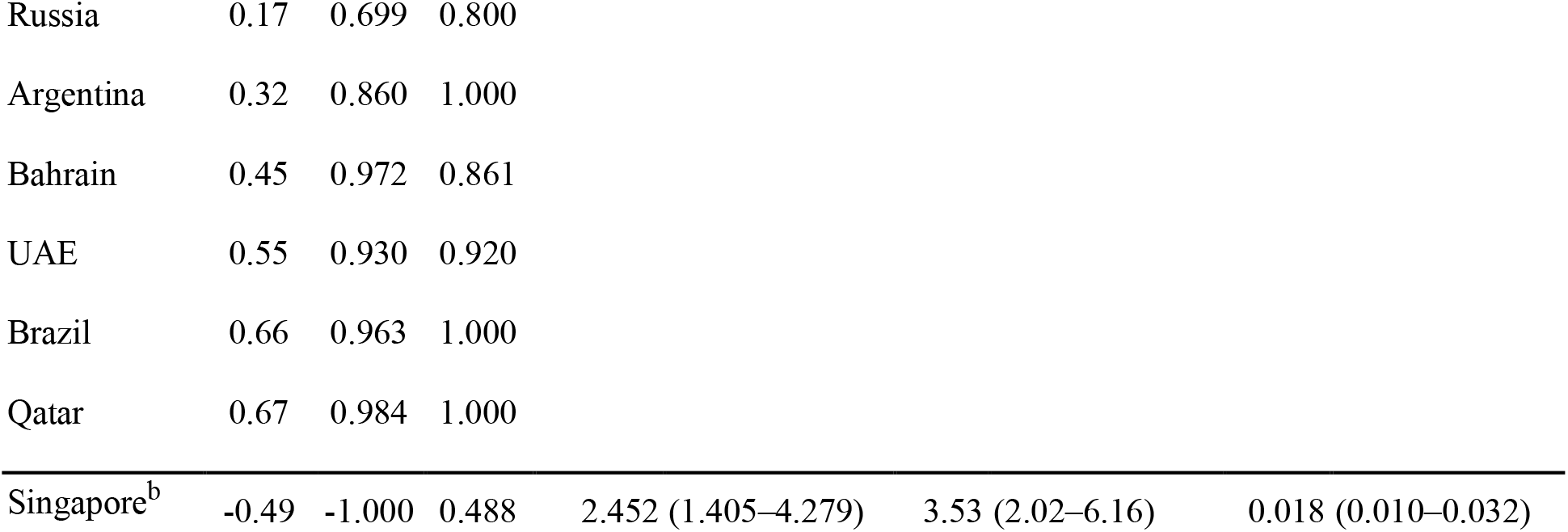
Parameters of the COVID-19 pandemic in representative countries.

## Discussion

So far, TIR has only been inferred from local surveys using PCR tests or antibody tests [4–7]. The Infection Fatality Rate (IFR) is unknown when TIR is not known. The mortality rate per population has been frequently used instead [8], although this is a product of TIR and IFR and is completely dependent on TIR. Another similar index, case fatality rate would be greatly influenced by the way of selecting cases [9], specifically the policy on conducting PCR tests. Estimating TIR and IFR by our method will contribute to consider the public health strategy.

The present method has several limitations. First, we cannot estimate TIR or IFR for countries in which the pandemic is still expanding. Second, the theoretical ground for our key hypothesis (negative correlation between ER and IR) may be obscure. However, the extremely high correlation found in the countries where the pandemic is relatively well controlled supports the validity of our estimate.

Comparing the present results with local surveys is interesting. Among European countries, a population-based study was conducted in Geneva, Switzerland and revealed 5 to 10% IR [4], which is much higher than our estimate of TIR for the whole Switzerland, 0.52%. Infected persons at the latest survey (June 13th) comprised 0.36% of the whole population of Switzerland (5.3% ER and 6.8% IR). When we divide the examined population into those up to the 3rd survey (May 16th) and those newly added between the 3rd and the latest surveys, each comprising 3.9% and 1.4% of the whole population, respectively, the IR was 9.0% for the former and 0.4% for the latter. The fact that the positive PCR rate is much lower for the recently expanded population supports our cardinal hypothesis. The 5–10% IR documented by the antibody survey [4] is inconsistent with the above figures. This may be due to the exceptionally high prevalence of COVID-19 in Geneva within Switzerland [10].

The reported IR in Tokyo, Japan, the most-densely infected region within Japan, varies from 0.10% (antibody tests) [7] to 6% (PCR tests) [5]. Our estimate of TIR in the whole Japan is 0.10% although the CI is wide (0.04–0.30) because of the inaccurate extrapolation due to the low ER. Local antibody survey may not give sufficient information for the whole country. Furthermore, the reliability of the antibody test may not be always guaranteed [11,12]. In this regard, our methods would have a certain role.

Estimated IFR also took on a wide range of values, up to 10%: generally high in the Western countries. Singapore achieved a straight-line for the latter 3 points, and the regression analysis was conducted for these points. The estimated IFR was extremely low, 0.018%. In Singapore, a gene mutation causing the attenuation of the viral virulence has been discovered [13]. Complete containment might be achieved by social distancing [2], which would however cause a serious economic damage. Herd immunity would be a long way considering the estimated low TIR. Attenuation of the viral virulence is another promising way to make the pandemic less damaging [14]. This is achieved in animals by natural selection [15], but in humans the prevention of in-hospital infection, especially from severe cases, would be the key.

## Data Availability

Prof. Masahiro Sonoo, the corresponding author has the availability of all data referred to in the manuscript.

## Figure Legend

**Supplementary Table 1.** Parameters of the COVID-19 pandemic in 64 investigated countries.

| Country or Region | slope | r | CR | TIR (%) (95%CI) | TICR (95%CI) | IFR (%) (95%CI) |
| --- | --- | --- | --- | --- | --- | --- |
| New Zealand | -1.01 | -0.999 | 0.000 | 0.029 (0.020–0.042) | 0.97 (0.68–1.39) | 1.503 (1.049–2.155) |
| Iceland | -0.97 | -1.000 | 0.004 | 0.560 (0.553–0.567) | 1.06 (1.04–1.07) | 0.523 (0.517–0.530) |
| Thailand | -0.94 | -1.000 | 0.030 | 0.006 (0.006–0.007) | 1.36 (1.25–1.48) | 1.360 (1.254–1.476) |
| Australia | -0.93 | -1.000 | 0.025 | 0.034 (0.034–0.035) | 1.19 (1.17–1.21) | 1.168 (1.147–1.189) |
| Luxembourg | -0.91 | -1.000 | 0.024 | 0.774 (0.639–0.937) | 1.19 (0.98–1.44) | 2.273 (1.877–2.753) |
| Slovenia | -0.88 | -1.000 | 0.015 | 0.105 (0.085–0.130) | 1.46 (1.18–1.81) | 4.977 (4.018–6.166) |
| Switzerland | -0.88 | -0.999 | 0.014 | 0.518 (0.373–0.720) | 1.44 (1.04–2.00) | 4.324 (3.112–6.009) |
| Hong Kong | -0.87 | -1.000 | 0.039 | 0.023 (0.021–0.026) | 1.58 (1.41–1.77) | 0.228 (0.204–0.256) |
| Croatia | -0.86 | -0.999 | 0.015 | 0.097 (0.062–0.152) | 1.77 (1.14–2.76) | 2.681 (1.719–4.183) |
| Austria | -0.86 | -1.000 | 0.040 | 0.282 (0.277–0.288) | 1.49 (1.46–1.52) | 2.664 (2.615–2.714) |
| Greece | -0.85 | -1.000 | 0.082 | 0.053 (0.044–0.064) | 1.78 (1.48–2.13) | 3.309 (2.759–3.968) |
| Lithuania | -0.84 | -0.998 | 0.150 | 0.086 (0.058–0.128) | 1.33 (0.90–1.98) | 3.184 (2.145–4.727) |
| Belgium | -0.83 | -1.000 | 0.159 | 0.770 (0.702–0.844) | 1.49 (1.36–1.63) | 10.821 (9.870–11.862) |
| Israel | -0.83 | -0.995 | 0.041 | 0.310 (0.152–0.632) | 1.50 (0.74–3.06) | 1.052 (0.516–2.147) |
| Spain | -0.83 | -1.000 | 0.056 | 0.944 (0.838–1.063) | 1.52 (1.35–1.71) | 6.147 (5.458–6.924) |
| S. Korea | -0.81 | -0.999 | 0.061 | 0.049 (0.033–0.075) | 2.10 (1.38–3.18) | 1.093 (0.720–1.660) |
| Italy | -0.80 | -0.999 | 0.094 | 0.667 (0.471–0.944) | 1.70 (1.20–2.41) | 8.506 (6.008–12.044) |
| Denmark | -0.79 | -1.000 | 0.144 | 0.320 (0.308–0.333) | 1.52 (1.46–1.58) | 3.218 (3.097–3.343) |
| Malaysia | -0.78 | -0.995 | 0.471 | 0.059 (0.021–0.165) | 2.27 (0.82–6.30) | 0.632 (0.227–1.756) |
| Germany | -0.78 | -0.999 | 0.083 | 0.431 (0.301–0.617) | 1.93 (1.35–2.76) | 2.456 (1.717–3.515) |
| Norway | -0.76 | -0.999 | 0.068 | 0.331 (0.256–0.429) | 2.08 (1.61–2.70) | 1.347 (1.040–1.745) |
| Estonia | -0.74 | -0.999 | 0.122 | 0.291 (0.213–0.397) | 1.96 (1.43–2.67) | 1.788 (1.311–2.438) |
| Serbia | -0.72 | -1.000 | 0.142 | 0.356 (0.327–0.389) | 2.54 (2.33–2.77) | 0.812 (0.745–0.886) |
| Morocco | -0.71 | -0.994 | 0.319 | 0.094 (0.023–0.382) | 3.96 (0.97–16.13) | 0.612 (0.151–2.490) |
| Ireland | -0.70 | -0.997 | 0.071 | 1.148 (0.720–1.829) | 2.24 (1.41–3.57) | 3.010 (1.889–4.797) |
| UK | -0.70 | -0.998 | 0.472 | 0.917 (0.602–1.395) | 2.11 (1.39–3.22) | 6.697 (4.400–10.193) |
| Netherlands | -0.68 | -0.997 | 0.159 | 0.931 (0.466–1.859) | 3.27 (1.64–6.53) | 3.799 (1.902–7.589) |
| Japan | -0.66 | -0.996 | 0.046 | 0.103 (0.035–0.302) | 7.52 (2.58–21.97) | 0.706 (0.242–2.063) |
| Czechia | -0.66 | -0.997 | 0.199 | 0.263 (0.149–0.465) | 2.82 (1.59–4.98) | 1.165 (0.659–2.061) |
| Hungary | -0.61 | -0.997 | 0.228 | 0.184 (0.096–0.355) | 4.38 (2.27–8.43) | 3.156 (1.638–6.081) |
| Portugal | -0.61 | -0.996 | 0.300 | 0.878 (0.569–1.355) | 2.46 (1.59–3.79) | 1.689 (1.094–2.606) |
| Finland | -0.54 | -0.991 | 0.217 | 0.585 (0.265–1.292) | 4.56 (2.07–10.08) | 1.006 (0.455–2.221) |
| Turkey | -0.54 | -0.995 | 0.233 | 1.079 (0.583–1.997) | 5.15 (2.78–9.53) | 0.527 (0.285–0.975) |
| Romania | -0.51 | -0.999 | 0.464 | 0.639 (0.489–0.836) | 5.59 (4.28–7.31) | 1.146 (0.877–1.498) |
| *USA<sup>a</sup>* | -0.47 | -0.999 | 0.646 | *2.599 (2.200–3.070)* | *4.01 (3.40–4.74)* | *1.367 (1.157–1.614)* |
| *Poland<sup>a</sup>* | -0.35 | -0.997 | 0.956 | *0.732 (0.520–1.029)* | *9.42 (6.70–13.25)* | *0.450 (0.320–0.634)* |
| Iran | -0.33 | -0.989 | 0.689 |  |  |  |
| Philippines | -0.32 | -0.941 | 0.972 |  |  |  |
| Moldova | -0.30 | -0.926 | 0.861 |  |  |  |
| Canada | -0.29 | -0.933 | 0.571 |  |  |  |
| *Indonesia<sup>a</sup>* | -0.26 | -0.995 | 0.920 | *1.395 (0.783–2.487)* | *99.66 (55.90–177.68)* | *0.056 (0.031–0.100)* |
| Ukraine | -0.25 | -0.986 | 0.803 |  |  |  |
| Ecuador | -0.19 | -0.939 | 0.194 |  |  |  |
| Dominican Republic | -0.18 | -0.989 | 0.953 |  |  |  |
| Sweden | -0.18 | -0.918 | 0.816 |  |  |  |
| Singapore | -0.16 | -0.620 | 0.488 |  |  |  |
| Panama | 0.00 | 0.004 | 1.000 |  |  |  |
| Belarus | 0.05 | 0.383 | 0.972 |  |  |  |
| India | 0.09 | 0.690 | 1.000 |  |  |  |
| Mexico | 0.13 | 0.935 | 1.000 |  |  |  |
| Russia | 0.17 | 0.699 | 0.800 |  |  |  |
| Saudi Arabia | 0.19 | 0.964 | 0.815 |  |  |  |
| Peru | 0.20 | 0.996 | 1.000 |  |  |  |
| Colombia | 0.25 | 0.983 | 1.000 |  |  |  |
| Argentina | 0.32 | 0.860 | 1.000 |  |  |  |
| Pakistan | 0.32 | 0.975 | 0.923 |  |  |  |
| Iraq | 0.40 | 0.762 | 1.000 |  |  |  |
| South Africa | 0.40 | 0.944 | 1.000 |  |  |  |
| Bahrain | 0.45 | 0.972 | 0.861 |  |  |  |
| Chile | 0.51 | 0.989 | 1.000 |  |  |  |
| Azerbaijan | 0.54 | 0.821 | 1.000 |  |  |  |
| UAE | 0.55 | 0.930 | 0.920 |  |  |  |
| Brazil | 0.66 | 0.963 | 1.000 |  |  |  |
| Qatar | 0.67 | 0.984 | 1.000 |  |  |  |
| Singapore<sup>b</sup> | -0.49 | -1.000 | 0.488 | 2.452 (1.405–4.279) | 3.53 (2.02–6.16) | 0.018 (0.010–0.032) |

